## Supplementary Information for "Congenital disorder of glycosylation caused by starting site-specific variant in syntaxin-5"

**Contents:**

- Supplementary Figures 1–18
- Supplementary Tables 1, 5 and 6
- Supplementary References

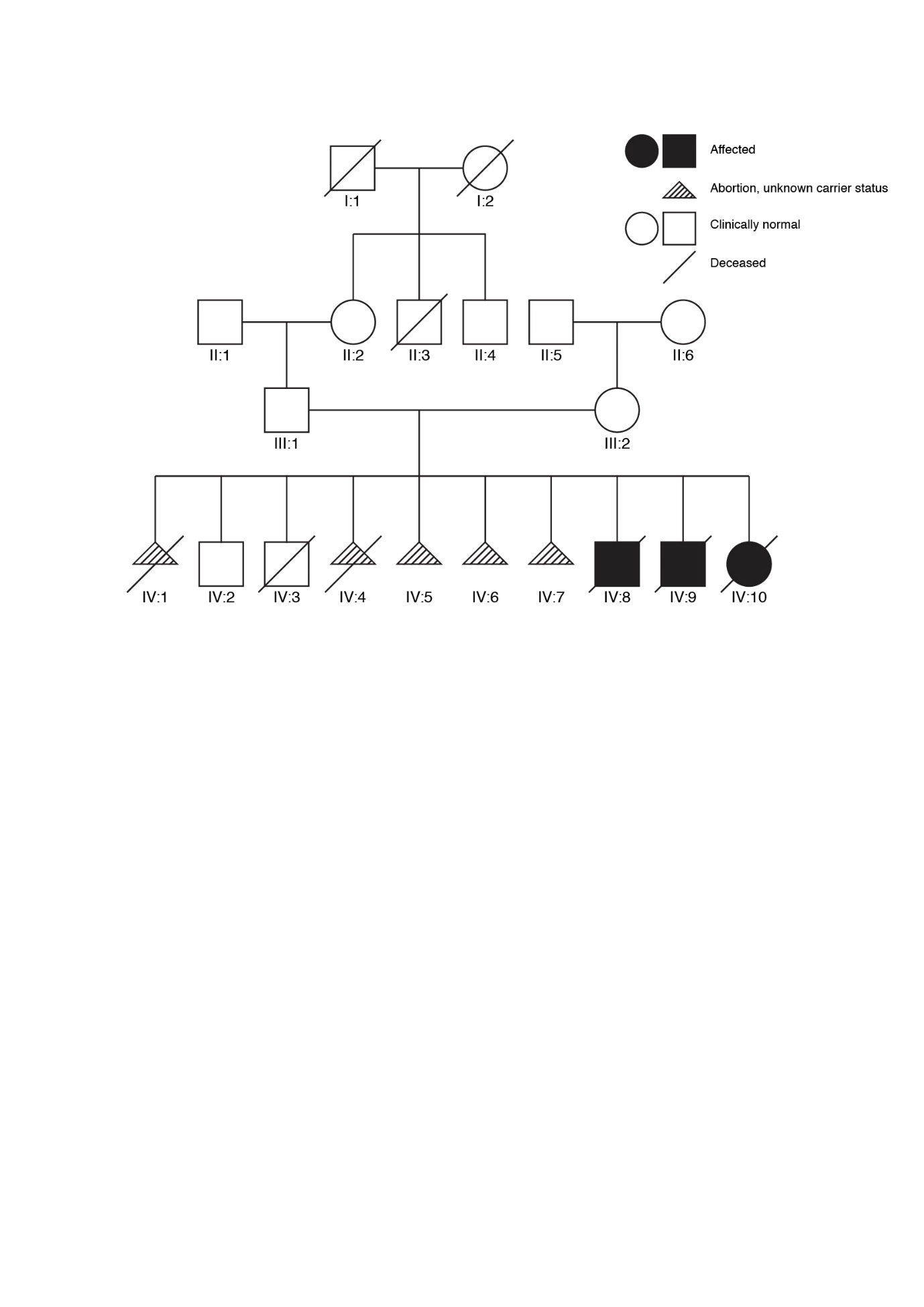

**Supplementary Figure 1. Family pedigree of Stx5M55V patients.**

Family pedigree of Stx5M55V patients. Black filled squares and circles indicate affected patients. Empty squares and circles indicate clinically normal individuals. Triangles with diagonal stripes indicate aborted individuals or individuals with unknown carrier status. The diagonal line indicates deceased individuals. Informed consent to carry out the research and publish the results was obtained from the family of investigated individuals.

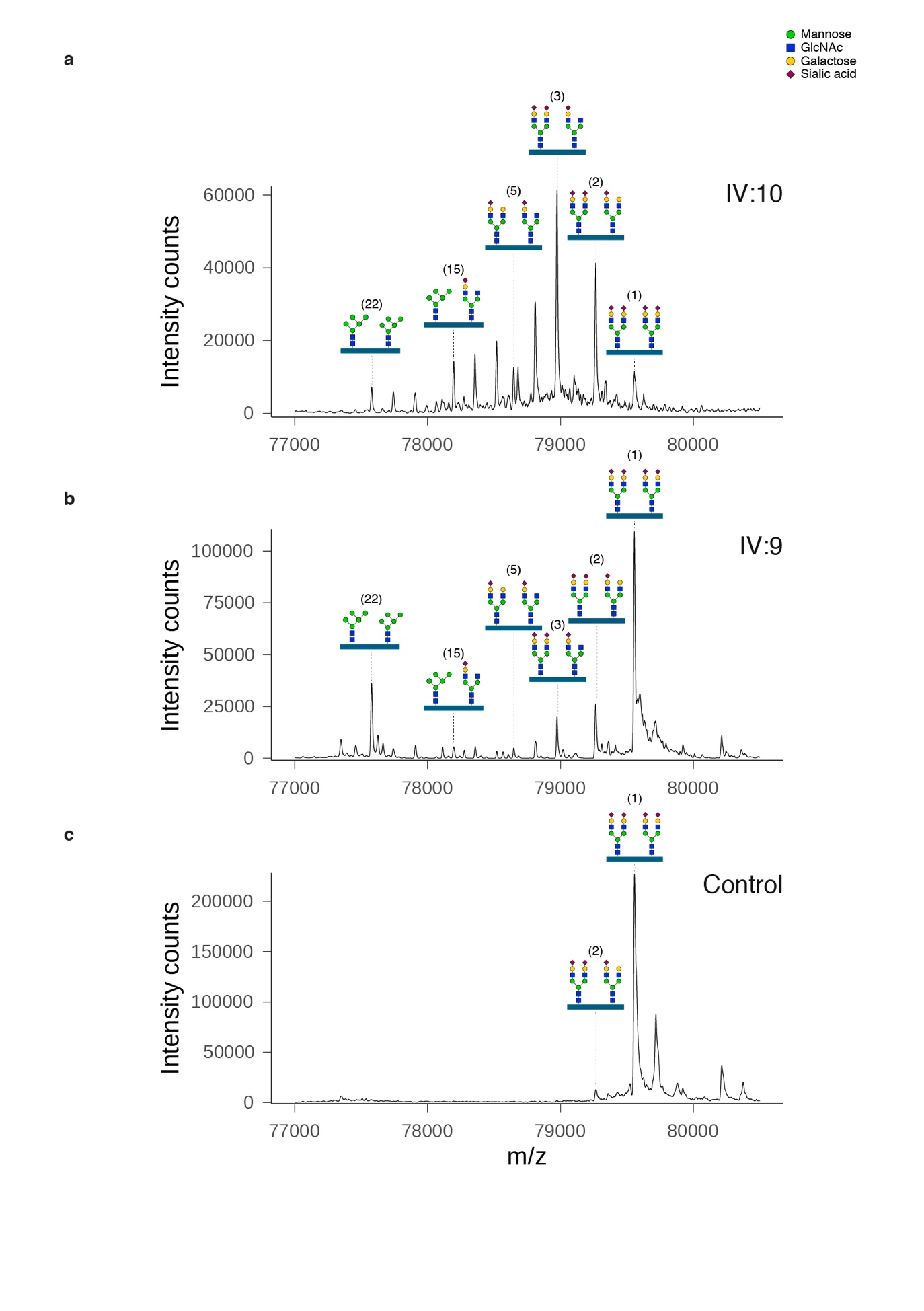
**Supplementary Figure 2. Glycan structural changes in STX5M55V patients by intact transferrin mass spectrometry.**

The nanochip-C8 QTOF mass spectra of enriched intact serum transferrin of Stx5M55V patient IV:10 (a), patient IV:9 (b) and healthy control (c) are shown. Annotation of all transferrin glycoforms is shown in Supplementary Table 2.

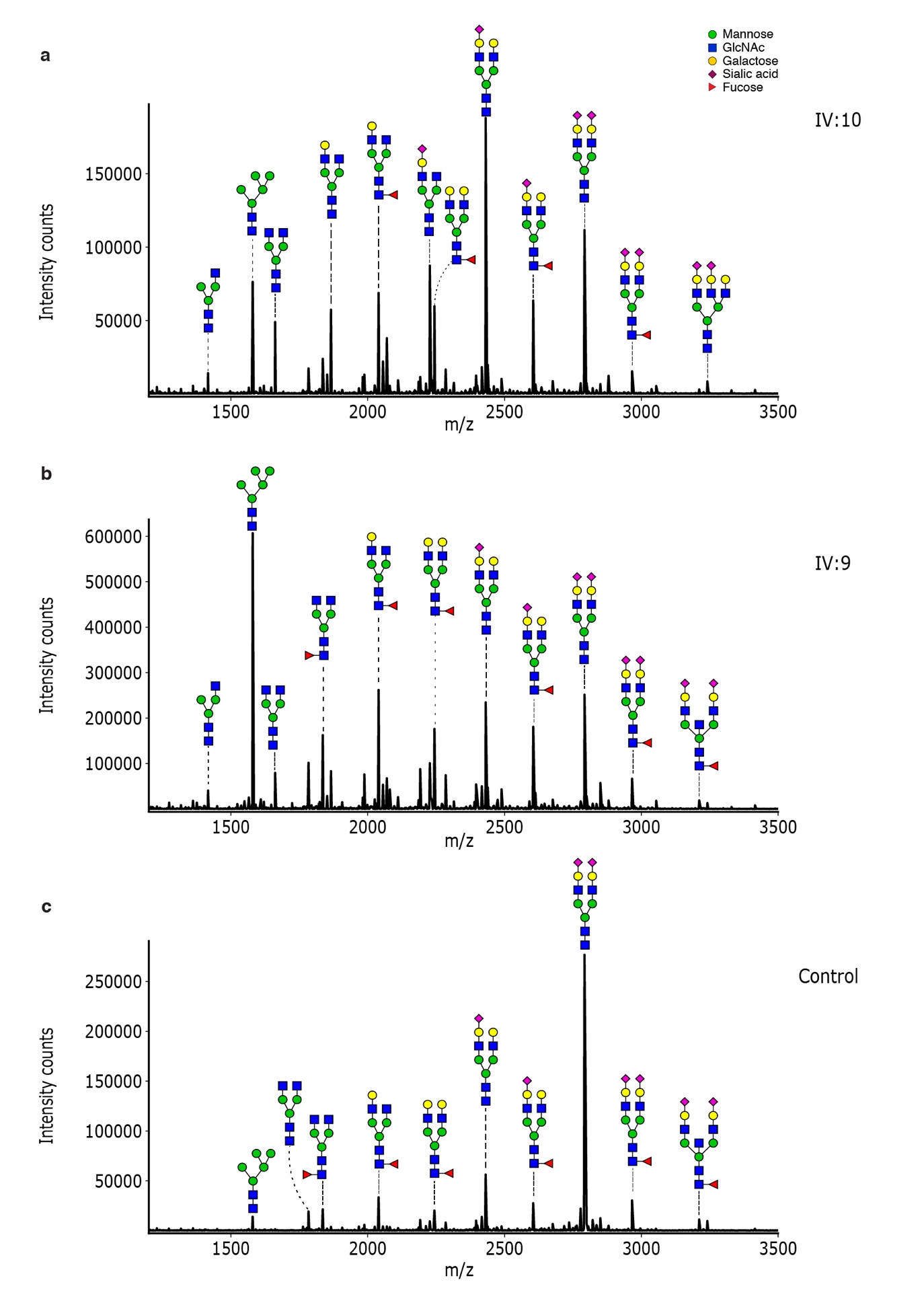

**Supplementary Figure 3. Glycan structural changes in STX5M55V patients by MALDI-TOF mass spectrometry of total plasma protein derived N-glycans.**

The MALDI-TOF mass spectra of all plasma N-glycans of Stx5M55V patient IV:10 (a), patient IV:9 (b) and healthy control (c) are shown. Annotation of all glycan structures is shown in Supplementary Table 3.

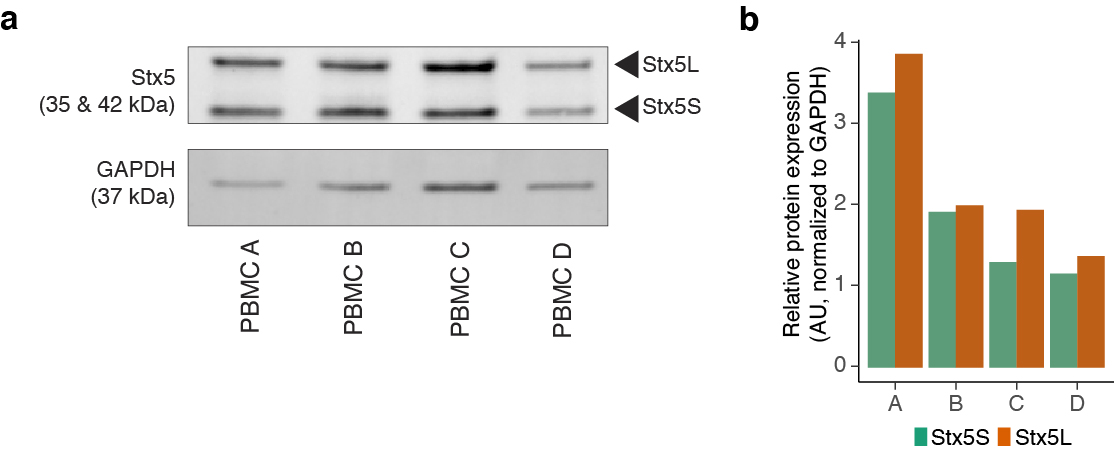
**Supplementary Figure 4. Interindividual variation in Stx5 expression.**

(a) Immunoblot for Stx5 on lysates of peripheral blood mononuclear cells (PBMCs). PBMCs were isolated from four healthy donors and analyzed side-by-side on a single Western blot for Stx5 to enable direct comparison. GAPDH, loading control.

(b) Quantification of (a). AU: arbitrary units.

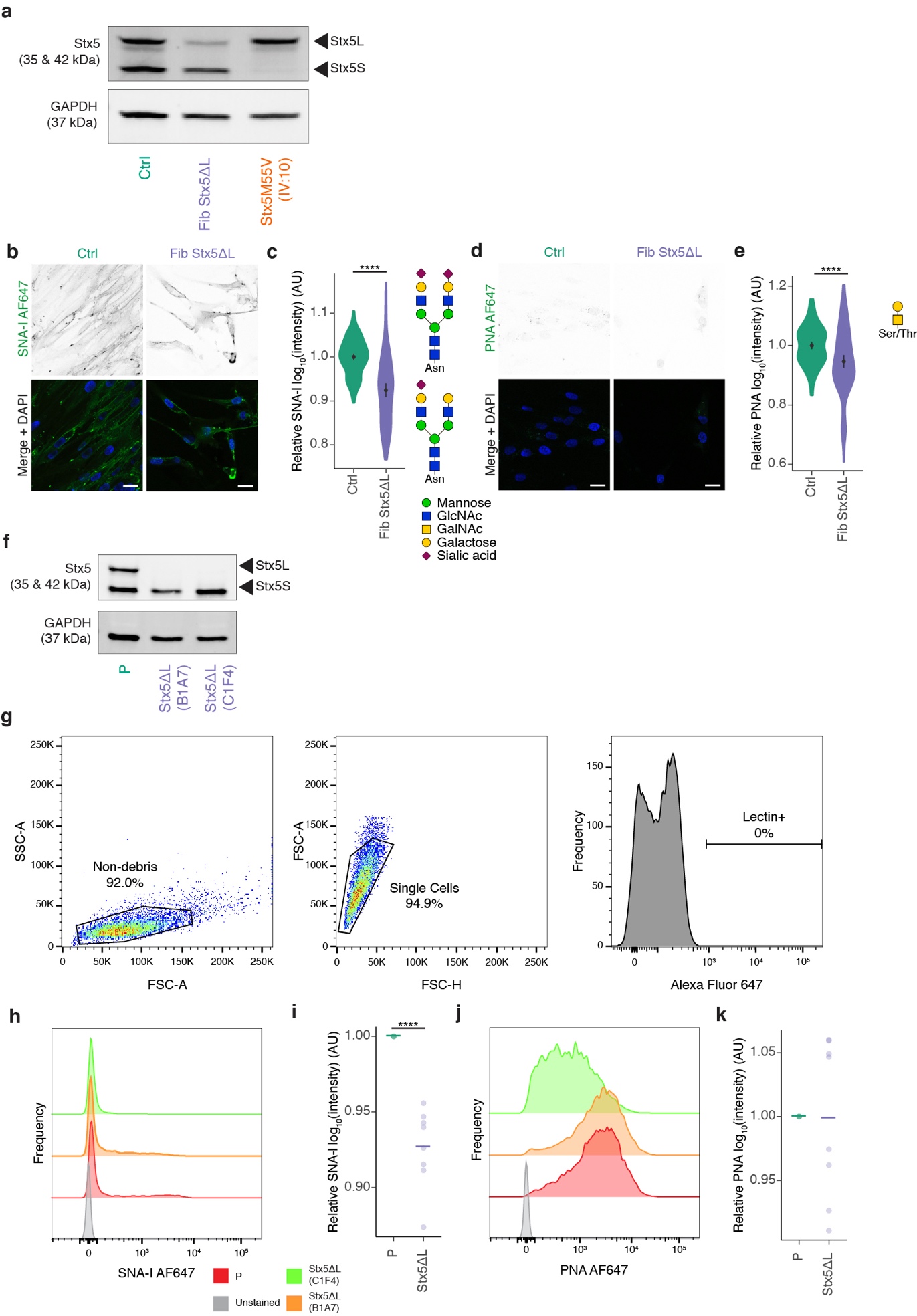
**Supplementary Figure 5. Stx5L-lacking cell line generation and lectin stainings.**

(a) Immunoblot for Stx5 of cell lysates of primary human dermal fibroblasts of a healthy donor (green, Ctrl), CRISPR-engineered Stx5L-lacking fibroblasts (blue, Fib Stx5ΔL) and fibroblasts from Stx5M55V patients (orange, Stx5M55V). GAPDH, loading control. The band corresponding to Stx5L in Fib Stx5ΔL shows residual Stx5ΔL expression due to incomplete CRISPR/Cas9 knockout. The Western blotting was performed twice to illustrate the deletions of Stx5S in Stx5M55V patients and Stx5L in Stx5L-lacking fibroblasts, which were confirmed by Sanger sequencing.

(b) Fibroblasts of healthy donors (green, Ctrl) or Stx5ΔL (blue, Fib Stx5ΔL) were probed with SNA-I lectin (green in merge). Representative confocal micrographs. Scalebars, 25 µm. DAPI in blue.

(c) Quantification of (b). All data were log_10_-transformed and then normalized to the healthy donor. AU: arbitrary units. N = 134 (Ctrl) and 107 (Stx5ΔL) from two independent cell lines repeated twice. Mean ± 95%CI. Unpaired two-sided Student’s t-test. ****: P = 2.3 × 10^-15^.

(d-e) Same as panels (b-c), but now for PNA lectin. N = 113 (Ctrl) and 111 (Stx5ΔL) from 2 independent cell lines repeated twice. Scalebars, 25 µm. Mean ± 95%CI. Unpaired two-sided Student’s t-test. ****: P = 2.91 × 10^-5^.

(f) Immunoblot for Stx5 of cell lysates of HeLa cells of parental (green, P) or two clonal Stx5L-lacking lines (blue, Stx5ΔL B1A7 and C1F4). GAPDH, loading control. The Western blotting was performed three times to illustrate the deletion of Stx5L in Stx5ΔL HeLa cell lines, which was confirmed by Sanger sequencing.

(g) FACS gating strategy for lectin stainings. Cell population shown is the unstained control. FSC-A: forward scatter area, SSC-A: side scatter area, FSC-H: forward scatter height.

(h) Representative FACS histogram of unstained cells (gray, Unstained) parental HeLa (red, P), or the two Stx5ΔL cell lines (orange, Stx5ΔL B1A7 and green Stx5ΔL C1F4), probed with SNA-I lectin conjugated to Alexa Fluor 647.

(i) Quantification of (h). Geometric means were taken and log_10_-transformed, then normalized to wildtype. 10,000 events analyzed per condition per experiment from 4 independent experiments with 2 Stx5ΔL lines. Average (bar) and all experimental averages plotted (dots). Unpaired two-sided Student’s t-test. ****: P = 1.0 × 10^-4^.

(j-k) Same as panels (h-i), but now for PNA lectin. 10,000 events analyzed per condition per experiment from 4 independent experiments with 2 Stx5ΔL lines. Average (bar) and all experimental averages plotted (dots). Not significant; unpaired two-sided Student’s t-test.

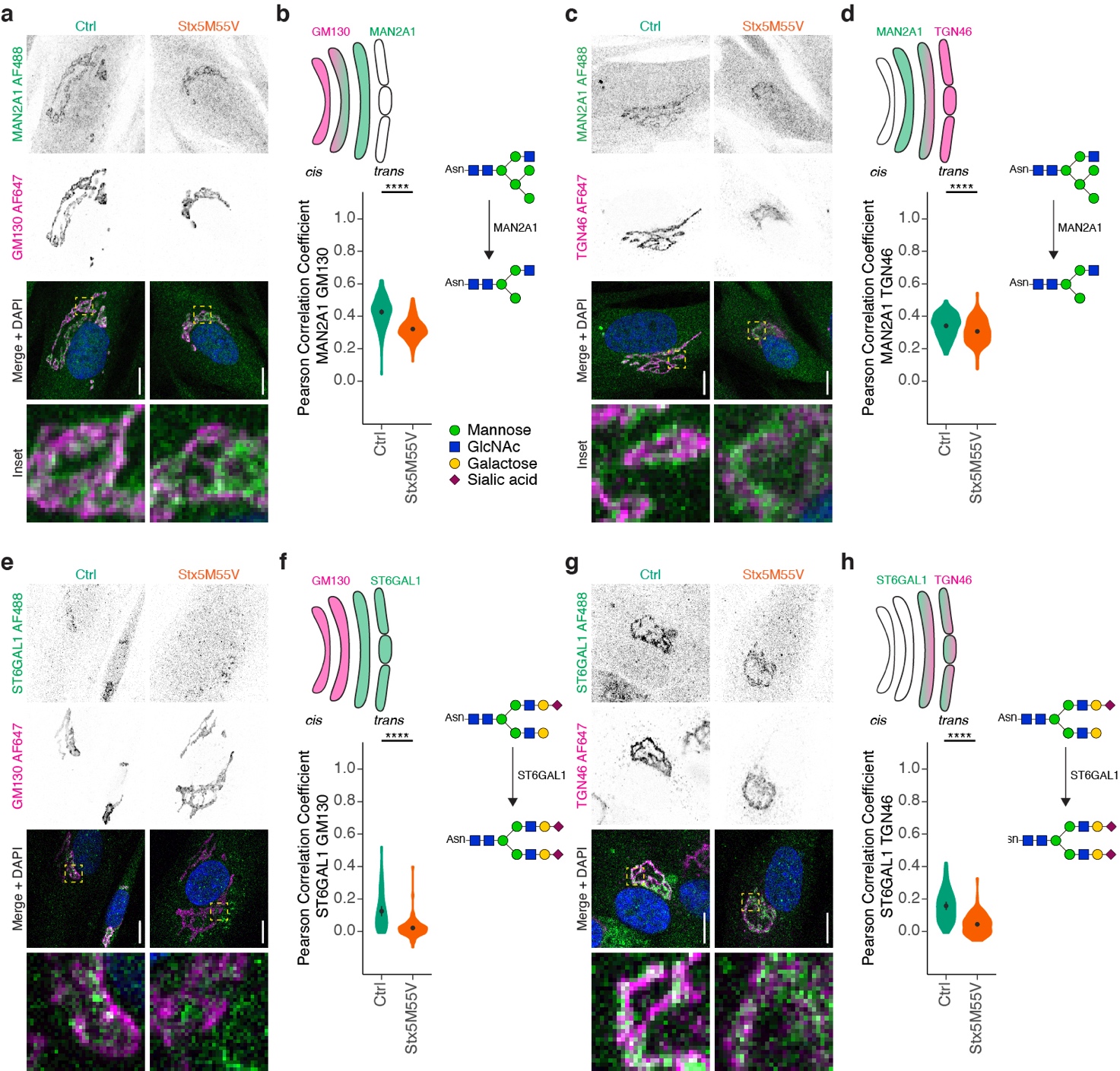

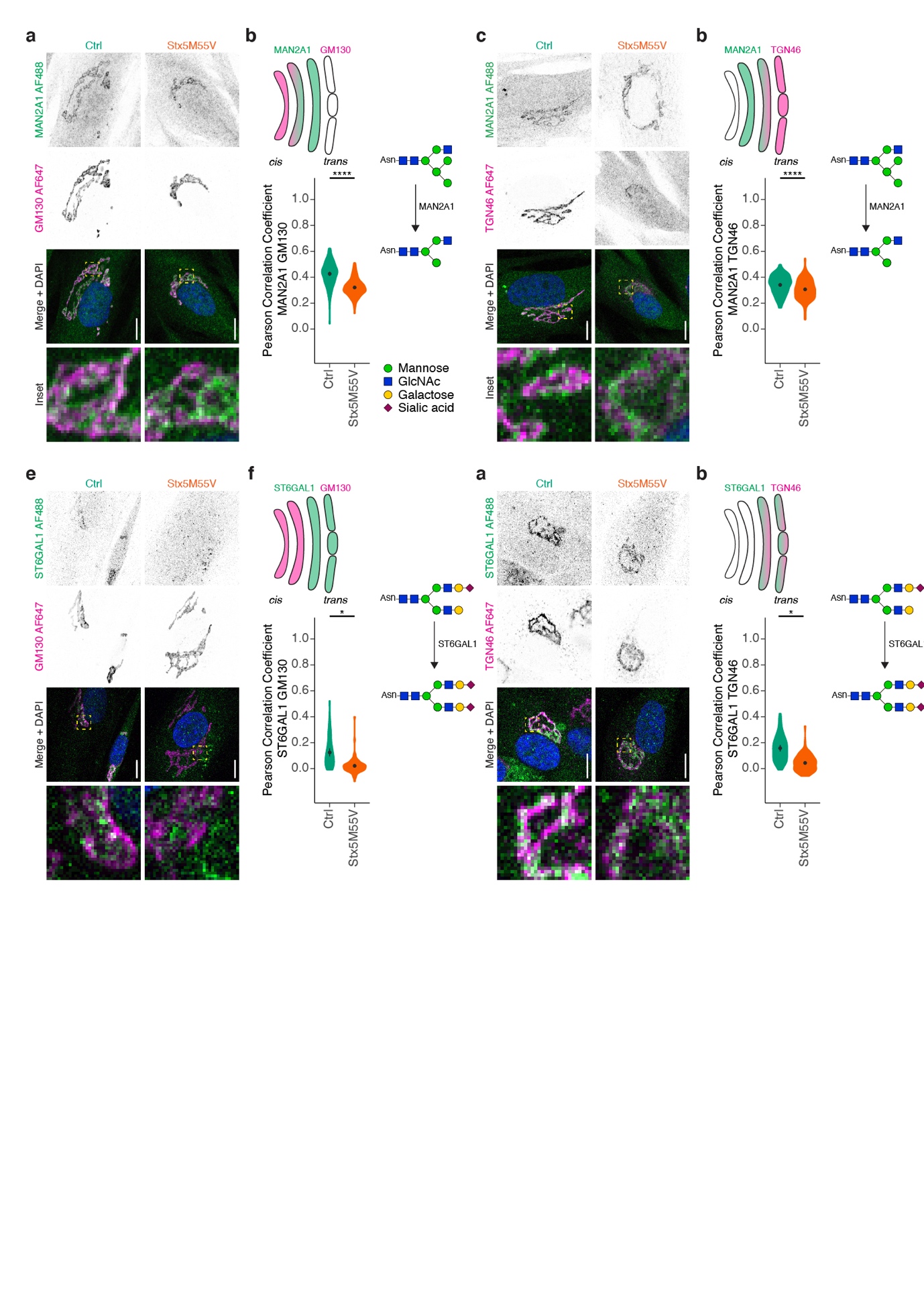

**Supplementary Figure 6. Glycosylation enzymes mislocalize in Stx5M55V patient fibroblasts.**

(a) Immunofluorescence microscopy of MAN2A1 (green in merge) and GM130 (magenta) in primary dermal fibroblasts of healthy donors (green, Ctrl) or Stx5M55V patients (orange, Stx5M55V). Representative confocal micrographs. Scalebars, 10 µm. DAPI in blue. N = 127 (Ctrl) and 143 (Stx5M55V) cells from 2 independent cell lines repeated twice.

(b) Pearson's correlations coefficients between MAN2A1 and GM130 of panel (a). Mean ± 95%CI. Unpaired two-sided Student’s t-test. ****: P = 6.4 × 10^-10^.

(c-d) Same as panels (a-b), but now for MAN2A1 (green) and TGN46 (magenta). N = 155 (Ctrl) and 171 (Stx5M55V) cells from 2 independent cell lines repeated twice. Scalebars, 10 µm. Mean ± 95%CI. Unpaired two-sided Student’s t-test. ****: P < 2.2 × 10^-16^.

(e-f) Same as panels (a-b), but now for ST6GAL1 (green) and GM130 (magenta). N = 56 (Ctrl) and 78 (Stx5M55V) cells from 2 independent cell lines repeated twice. Scalebars, 10 µm. Mean ± 95%CI. Unpaired two-sided Student’s t-test. ****: P = 4.4 × 10^-10^.

(g-h) Same as panels (a-b) but now for ST6GAL1 (green) and TGN46 (magenta). N = 57 (Ctrl) and 86 (Stx5M55V) cells from 2 independent cell lines repeated twice. Scalebars, 10 µm. Mean ± 95%CI. Unpaired two-sided Student’s t-test. ****: P = 2.1 × 10^-14^.

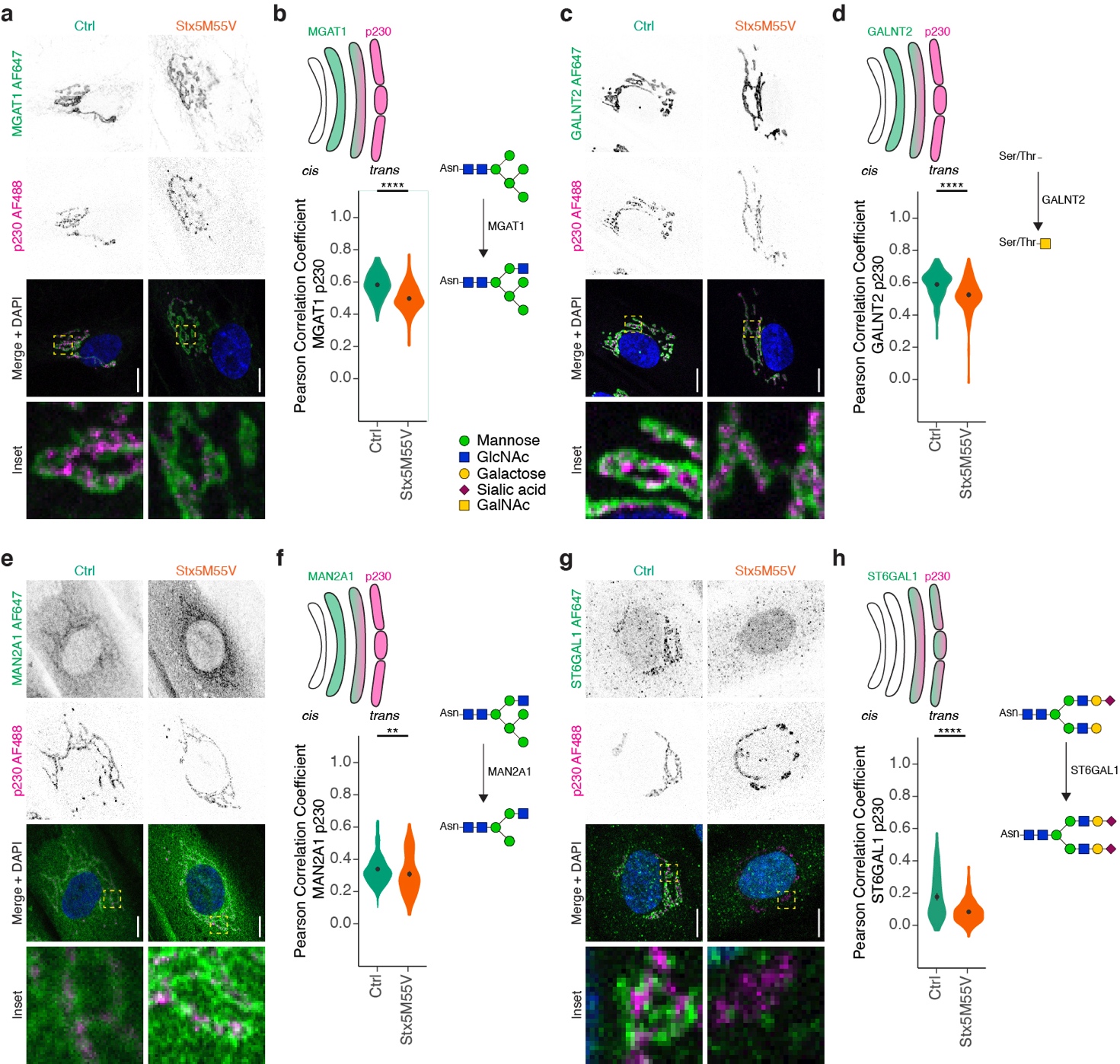

**Supplementary Figure 7. Glycosylation enzymes mislocalize in Stx5M55V patient fibroblasts.**

(a) Immunofluorescence microscopy of MGAT1 (green in merge) and p230 (magenta) in primary dermal fibroblasts of healthy donors (green, Ctrl) or Stx5M55V patients (orange, Stx5M55V). Representative confocal micrographs. Scalebars, 10 µm. DAPI in blue. N = 153 (Ctrl) and 147 (Stx5M55V) cells from 2 independent cell lines repeated twice.

(b) Pearson's correlations coefficients between MGAT1 and p230 of panel (a). Mean ± 95%CI. Unpaired two-sided Student’s t-test. ****: P < 2.2 × 10^-16^.

(c-d) Same as panels (a-b), but now for GALNT2 (green) and p230 (magenta). N = 166 (Ctrl) and 160 (Stx5M55V) cells from 2 independent cell lines repeated twice. Scalebars, 10 µm. Mean ± 95%CI. Unpaired two-sided Student’s t-test. ****: P = 6.4 × 10^-10^.

(e-f) Same as panels (a-b), but now for MAN2A1 (green) and p230 (magenta). N = 184 (Ctrl) and 75 (Stx5M55V) cells from 2 independent cell lines repeated twice. Scalebars, 10 µm. Mean ± 95%CI. Unpaired two-sided Student’s t-test. **: P = 0.0088.

(g-h) Same as panels (a-b) but now for ST6GAL1 (green) and p230 (magenta). N = 128 (Ctrl) and 149 (Stx5M55V) cells from 2 independent cell lines repeated twice. Scalebars, 10 µm. Mean ± 95%CI. Unpaired two-sided Student’s t-test. ****: P = 1.4 × 10^-12^.

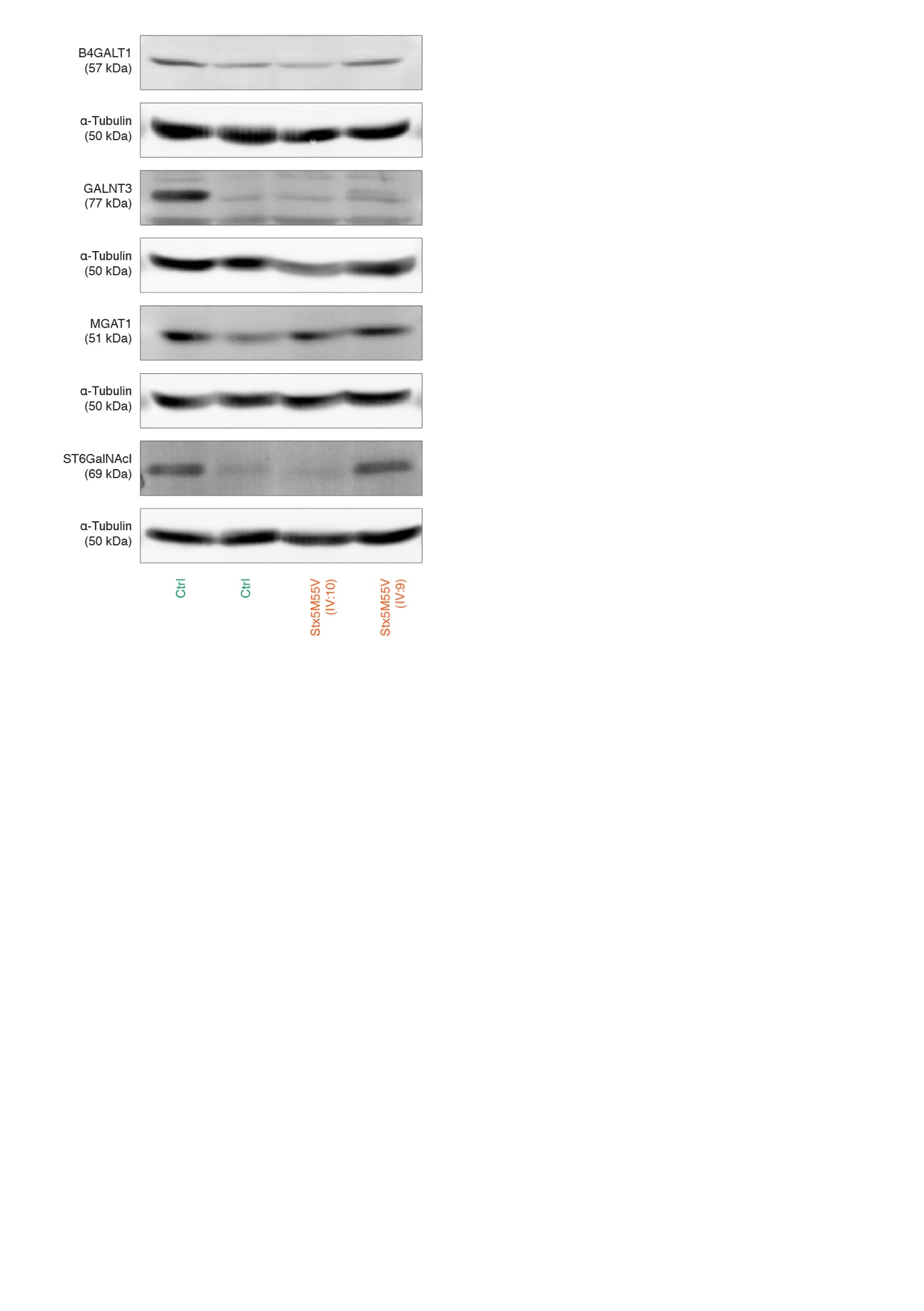

**Supplementary Figure 8. No consistent changes in expression of glycosyltransferases in healthy and patient fibroblasts.**

Immunoblots for glycosyltransferases of cell lysates of primary human dermal fibroblasts of healthy donors (green, Ctrl) or Stx5M55V patients (orange, Stx5M55V). Fibroblasts were derived from two unique healthy donors and two Stx5M55V patients and probed side-by-side on single Western blots to allow for direct comparison. The blots were first probed for the indicated glycosyltransferases and then for the loading control α-Tubulin.

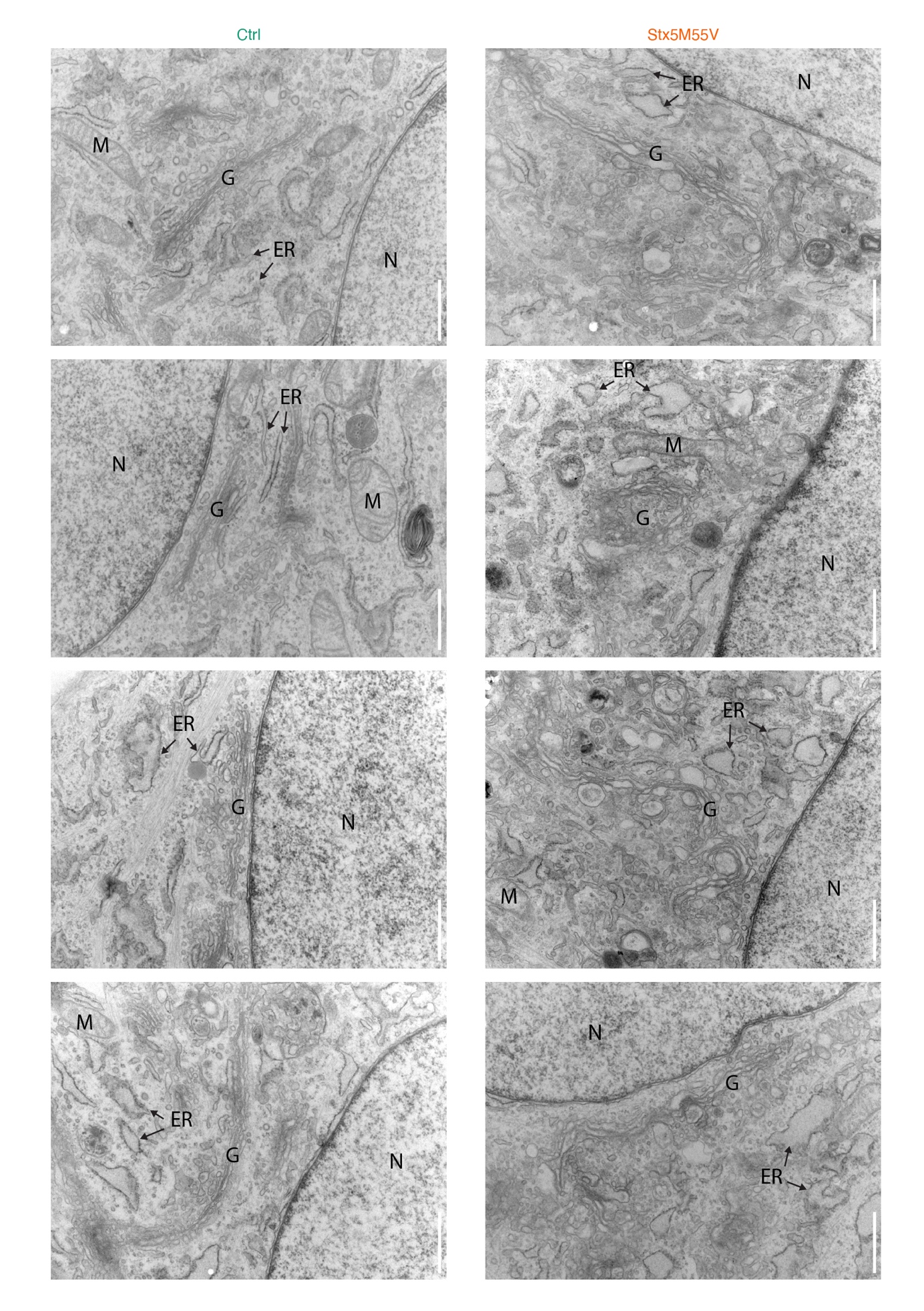
**Supplementary Figure 9. Gallery of representative transmission electron micrographs of healthy donor and Stx5M55V fibroblasts.** Images acquired in two independent repeats.

Scalebars, 1 µm. N, nucleus; G, Golgi apparatus; ER, endoplasmic reticulum; M, mitochondrion.

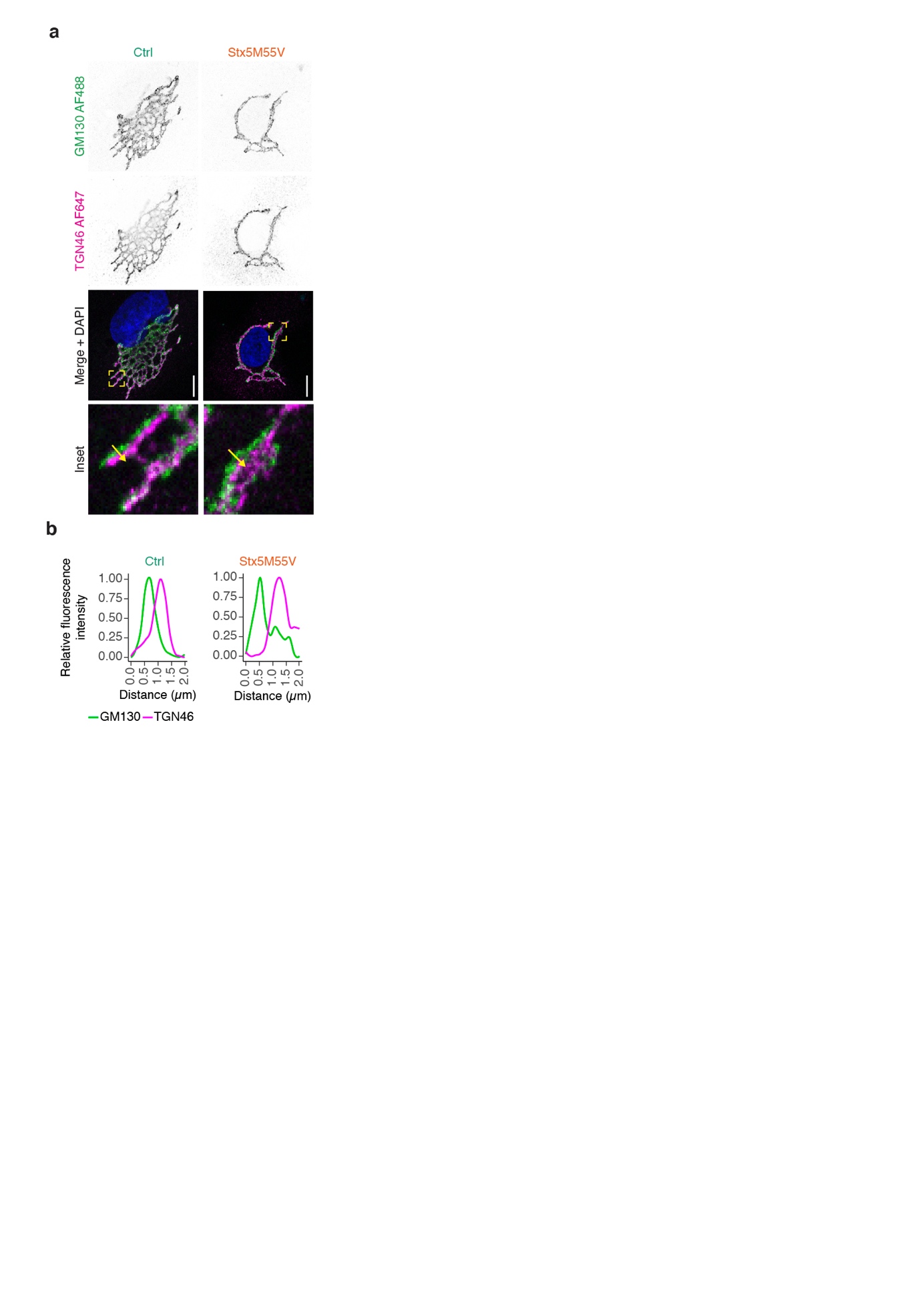

**Supplementary Figure 10. Golgi cisterna polarization is maintained in Stx5M55V patient fibroblasts.**

(a) Immunofluorescence microscopy of GM130 (green in merge) and TGN46 (magenta) in primary dermal fibroblasts of healthy donors (green, Ctrl) or Stx5M55V patients (orange, Stx5M55V). Representative confocal micrographs. Scalebars, 10 µm. DAPI in blue. The yellow arrows indicate the cross-sections used for the line plots in panel (b).

(b) Fluorescence intensity line plots from panel (a). Data for each marker was normalized to the maximum fluorescent intensity per cross-section.

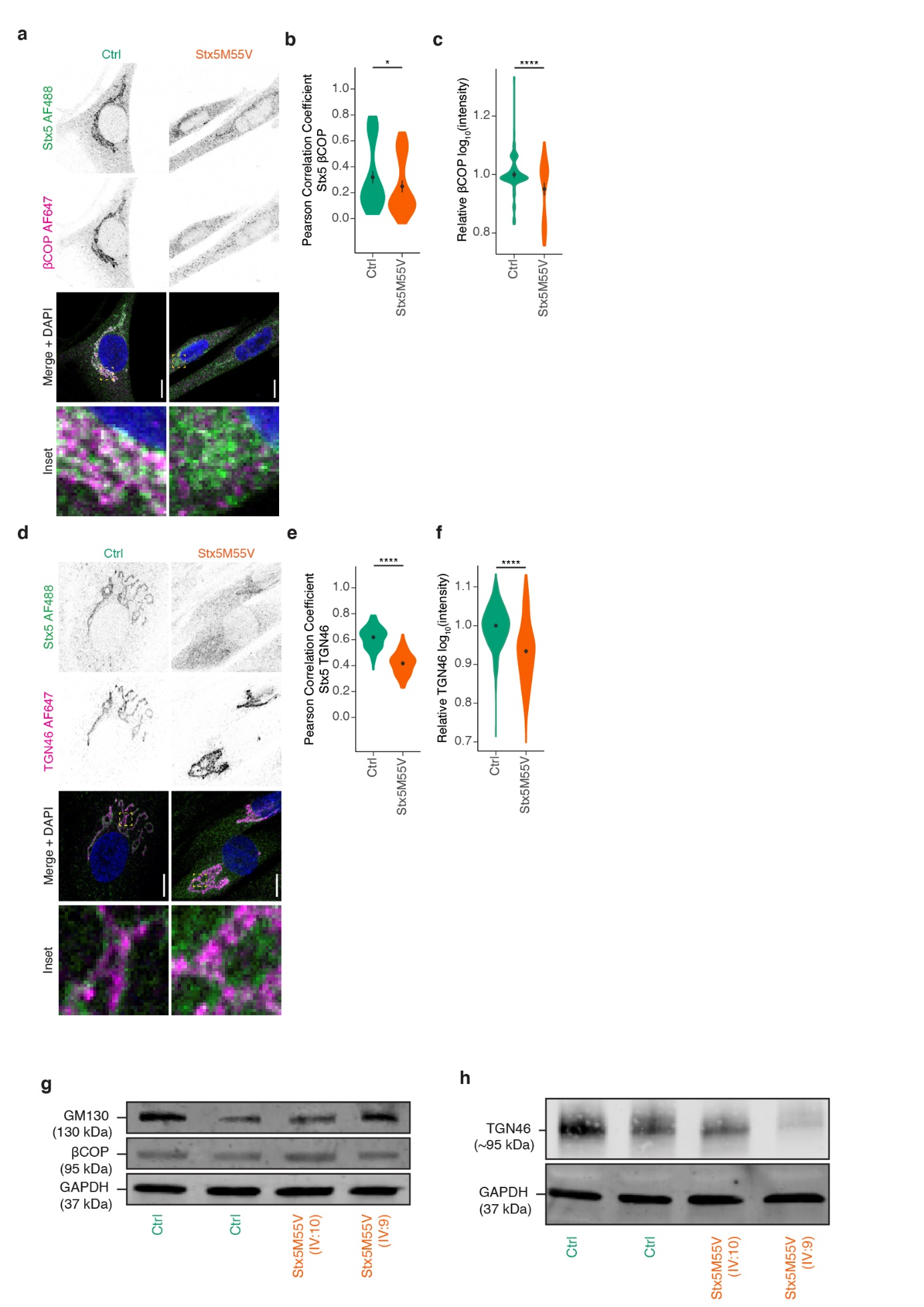

**Supplementary Figure 11. Reduced localization of Stx5 to trans-Golgi network in Stx5M55V patients.**

(a) Immunofluorescence microscopy of Stx5 (green in merge) and βCOP (magenta) in primary dermal fibroblasts of healthy donors (green, Ctrl) or Stx5M55V patients (orange, Stx5M55V). Representative confocal micrographs. Scalebars, 10 µm. DAPI in blue.

(b) Pearson's correlations coefficients between Stx5 and βCOP of panel (a). N = 109 (Ctrl) and 87 (Stx5M55V) cells from two independent cell lines repeated twice. Mean ±95%CI. Unpaired two-sided Student’s t-test. *: P = 0.034.

(c) Fluorescence intensities of βCOP from panel (a) relative to the healthy control. N = 109 (Ctrl) and 87 (Stx5M55V) cells from 2 independent cell lines repeated twice. Mean ±95%CI. Unpaired two-sided Student’s t-test. ****: P = 3.19 × 10^-5^.

(d-f) Same as panels (a-c), but now for Stx5 (green) and TGN46 (magenta). N = 128 (Ctrl) and 114 (Stx5M55V) cells from 2 independent cell lines repeated twice for colocalization, N = 822 (Ctrl) and 783 (Stx5M55V) cells from 2 independent cell lines repeated six times for intensity measurements. Scalebars, 10 µm. Mean ±95%CI. Unpaired two-sided Student’s t-test. ****: P < 2.2 × 10^-16^ for panels e and f.

(g-h) Immunoblot for GM130 and βCOP (g) or TGN46 (h) of the cells from panel A. Fibroblasts were derived from two unique healthy donors and two Stx5M55V patients and probed side-by-side on single Western blots to allow for direct comparison. The blots were first probed for the indicated proteins and then for the loading control GAPDH.

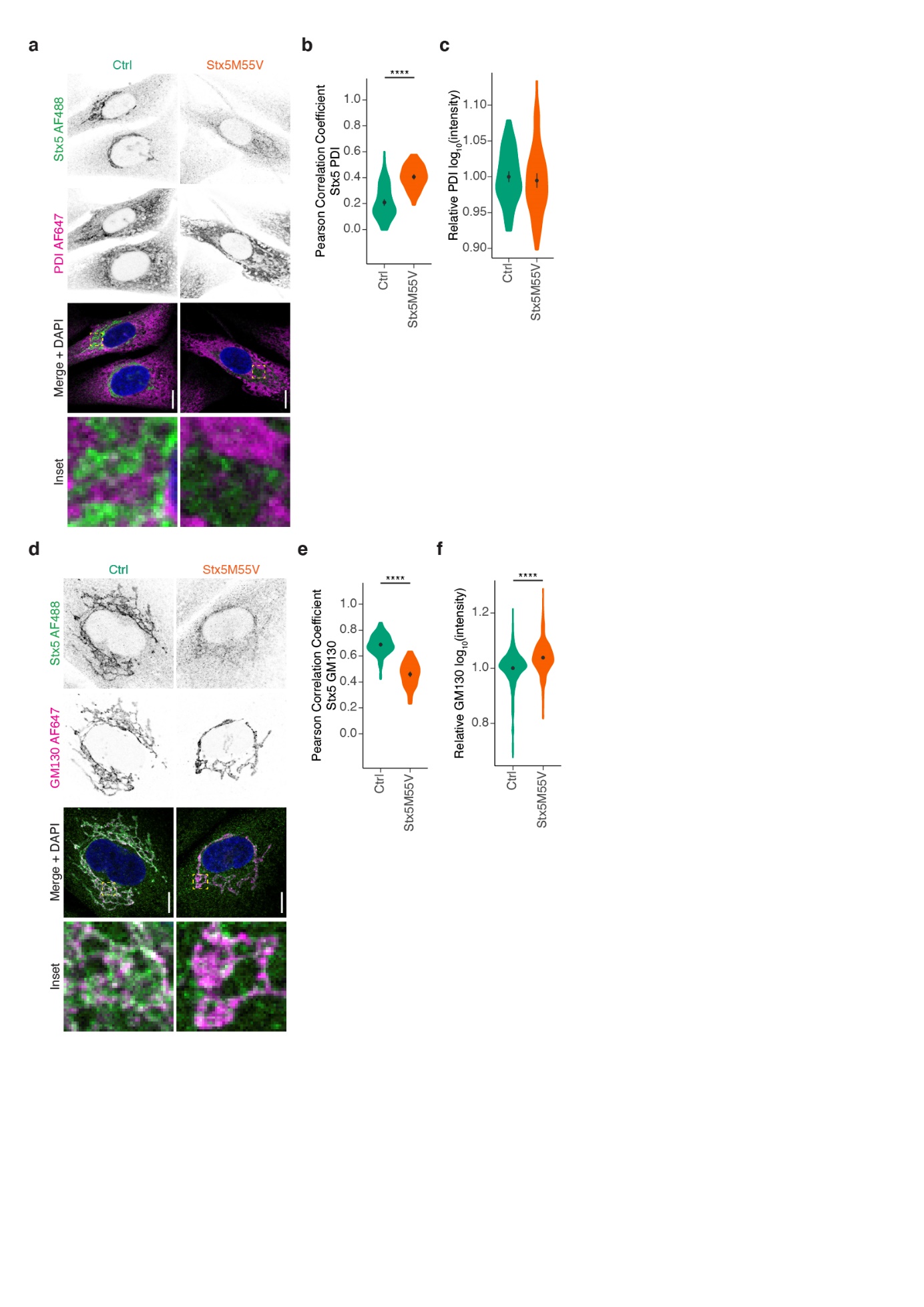

**Supplementary Figure 12. Stronger localization of Stx5 to ER in Stx5M55V patient fibroblasts.**

(a) Immunofluorescence microscopy of Stx5 (green in merge) and PDI (magenta) in primary dermal fibroblasts of healthy donors (green, Ctrl) or Stx5M55V patients (orange, Stx5M55V). Scalebars, 10 µm. DAPI in blue.

(b) Pearson's correlations coefficients between Stx5 and PDI of panel (a). N = 101 (Ctrl) and 76 (Stx5M55V) cells from 2 independent cell lines repeated twice. Mean ±95%CI. Unpaired two-sided Student’s t-test. ****: P < 2.2 × 10^-16^.

(c) Fluorescence intensity of PDI from panel (a) relative to healthy donor. N = 93 (Ctrl) and 84 (Stx5M55V) cells from 2 independent cell lines repeated twice. Mean ±95%CI. Not significant; unpaired two-sided Student’s t-test.

(d-f) Same as panels (a-c), but now for Stx5 (green) and GM130 (magenta). N = 105 (Ctrl) and 80 (Stx5M55V) cells from 2 independent experiments for colocalization, N = 415 (Ctrl) and 436 (Stx5M55V) cells from 2 independent cell lines repeated four times for intensity. Scalebars, 10 µm. Mean ±95%CI. Unpaired two-sided Student’s t-test. ****: P < 2.2 × 10^-16^ for panels e and f.

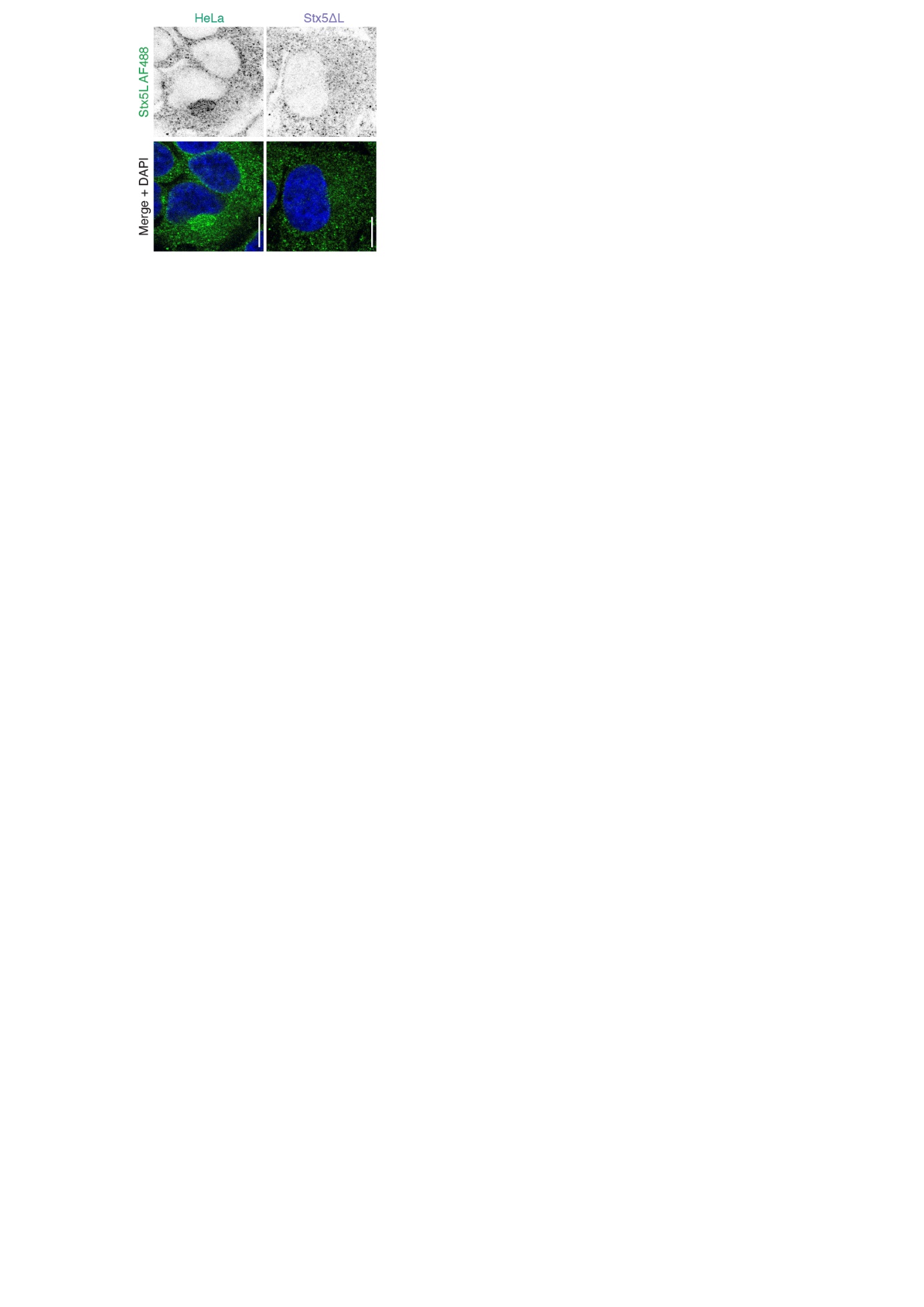
**Supplementary Figure 13. Stx5L-specific antibody validation.**

Immunofluorescence microscopy of Stx5L (green in merge) in parental HeLa (HeLa, green) or HeLa Stx5ΔL (Stx5ΔL, blue). Representative epifluorescence micrographs. Scalebars, 10 µm. DAPI in blue. Experiment conducted once to confirm the labeling specificity indicated by the manufacturer.

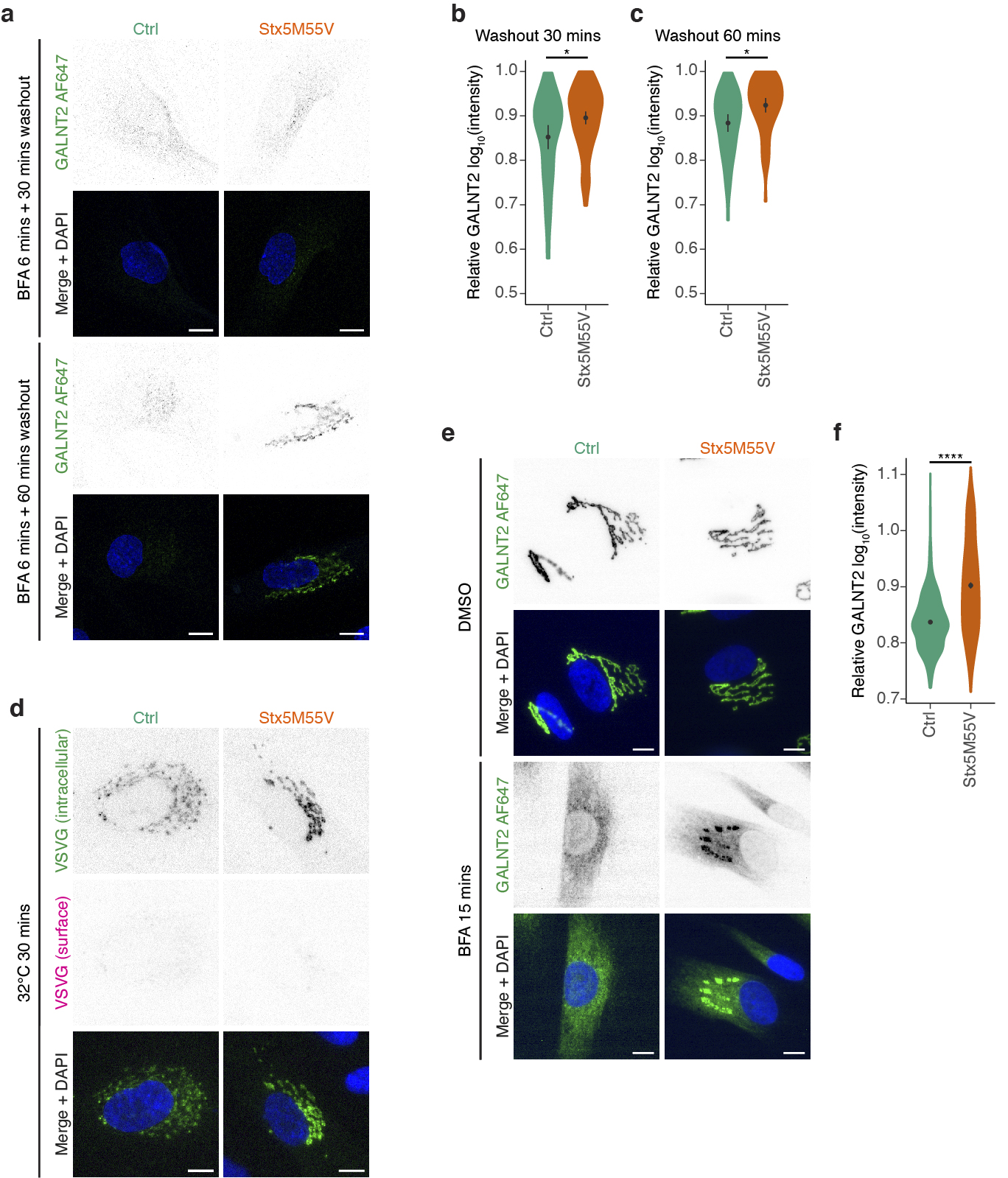
**Supplementary Figure 14. BFA and VSVG experiments in Stx5M55V fibroblasts.**

(a) Immunofluorescence microscopy of GALNT2 (green in merge) in primary human dermal fibroblasts of healthy donors (green, Ctrl) or Stx5M55V patients (orange, Stx5M55V) in the presence of Brefeldin A (BFA) for 6 min and washout for the indicated times. Representative confocal micrographs. Scalebars, 10 µm. DAPI in blue.

(b) Quantification of the 30 min timepoint from panel (a). N = 64 (Ctrl) and 119 (Stx5M55V) cells from 2 independent cell lines repeated twice. Mean ± 95%CI. Unpaired two-sided Student’s t-test and adjusted with Bonferroni correction for multiple testing. *: P = 0.048.

(c) Same as (b), but now for the 60 min timepoint. N = 68 (Ctrl) and 83 (Stx5M55V) cells from 2 independent cell lines repeated twice. Mean ± 95%CI. Unpaired two-sided Student’s t-test and adjusted with Bonferroni correction for multiple testing. *: P = 0.033.

(d) Immunofluorescence microscopy of VSVG-EGFP in primary human dermal fibroblasts of healthy donors (green, Ctrl) or Stx5M55V patients (orange, Stx5M55V) cultured overnight at 40°C and then 32°C for 30 mins. Representative confocal micrographs are shown. Scalebars, 10 µm. DAPI in blue.

(e) Same as panel (a), but now in the absence or presence of Brefeldin A (BFA) for 15 min. Representative epifluorescence micrographs are shown. Scalebars, 10 µm. DAPI in blue. N = 979 (Ctrl) and 943 (Stx5M55V) cells from 2 independent experiments.

(f) Relative maximum fluorescence intensities of GALNT2 from panel (e). All data were normalized to the DMSO condition (vehicle). Mean ± 95%CI. Unpaired two-sided Student’s t-test. ****: P < 2.2 × 10^-16^.

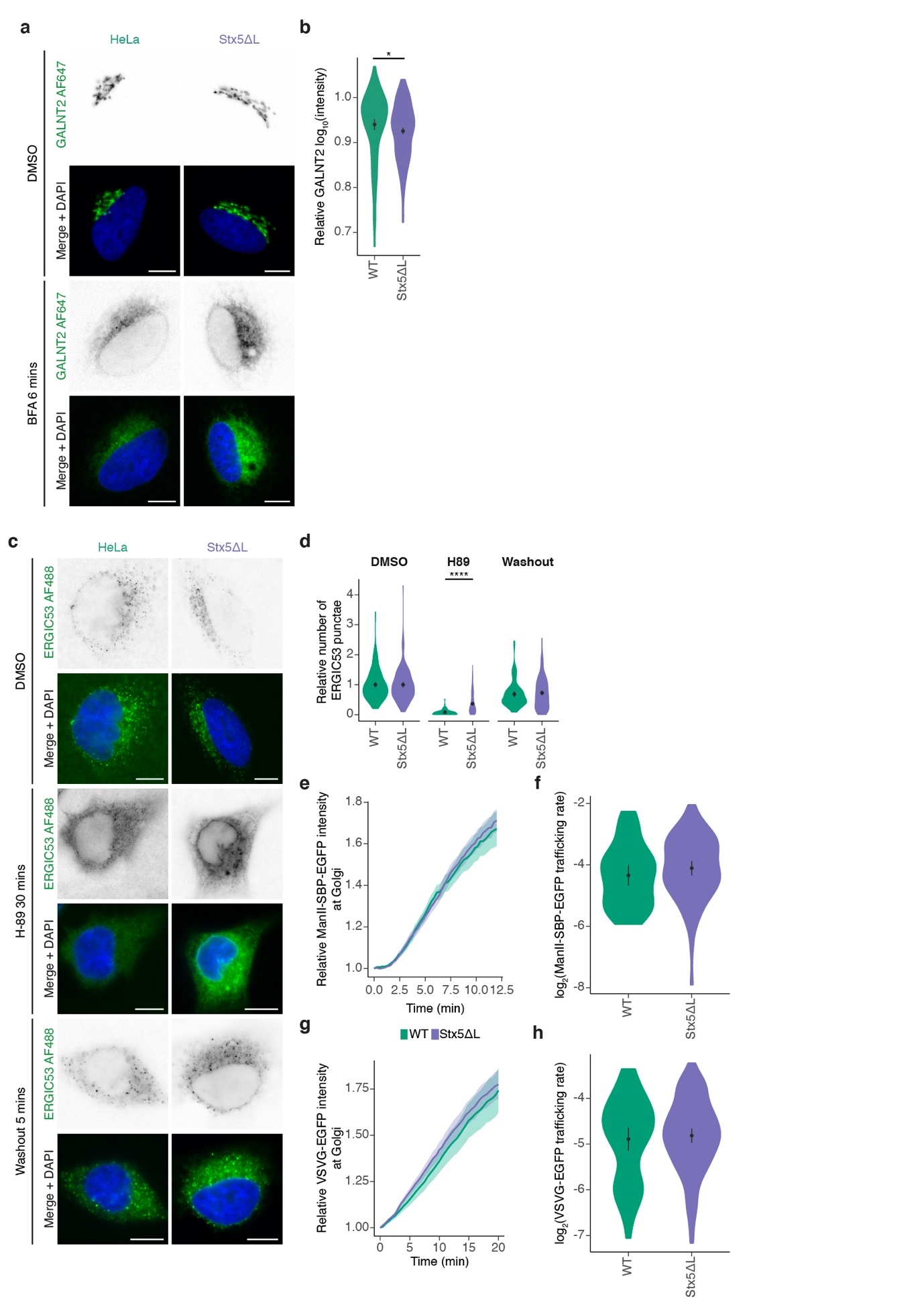

**Supplementary Figure 15. Loss of Stx5L does not affect anterograde ER-Golgi trafficking.**

(a) Immunofluorescence microscopy of GALNT2 (green in merge) in parental HeLa (WT, green) or HeLa Stx5ΔL (Stx5ΔL, blue) in the absence or presence of Brefeldin A (BFA) for 6 min. Representative epifluorescence micrographs. Scalebars, 10 µm. DAPI in blue.

(b) Relative maximum fluorescence intensities of GALNT2 from panel (a). All data were normalized to the DMSO condition (vehicle). N = 103 (WT) and 212 (Stx5ΔL) cells from 2 independent experiments. Mean ±95%CI. Unpaired two-sided Student’s t-test. *: P = 0.0335.

(c) Immunofluorescence microscopy of ERGIC53 (green in merge) in the absence or presence of H-89 and after washout of H-89 in parental HeLa (WT, green) or HeLa Stx5ΔL (Stx5ΔL, blue). H-89 inhibits export of cargo from ER exit sites via inhibition of protein kinase A, and causes redistribution of ERGIC53 to the ER, both of which are restored upon washout^1,2^. Representative epifluorescence micrographs. Scalebars, 10 µm. DAPI in blue.

(d) The relative number of ERGIC53 punctae of panel (c). All data were normalized to the average of the DMSO condition of each of the lines. N = 140 (WT DMSO), 90 (WT H-89), 92 (WT Washout), 169 (Stx5ΔL), 188 (Stx5ΔL H-89) and 167 (Stx5ΔL Washout) cells from 2 independent experiments. Mean ±95%CI. Mann-Whitney U test. ****: P < 2.2 × 10^-16^. Other comparisons not significant.

(e) Trafficking of ManII-SBP-EGFP in parental HeLa (WT, green) or HeLa Stx5ΔL (Stx5ΔL, blue) over time (n ~40 cells/condition). As part of the RUSH system^3^, ManII-SBP-EGFP is held at the ER in absence of biotin by interaction with a streptavidin-KDEL hook protein. After addition of biotin at t = 0, ManII-SBP-EGFP can exit the ER normally and traffic to the Golgi. ManII-SBP-EGFP intensity at the Golgi was measured, corrected to the total cellular intensity of ManII-SBP-EGFP per frame, and normalized to t = 0. See also supplementary movies 1 and 2. SBP, streptavidin binding protein. Shown is the mean ± error band (95%CI calculated for each timepoint and interpolated inbetween timepoints).

(f) Quantification of slope coefficients from panel (e) of the linear section (3-6 min). No differences between WT and Stx5ΔL were observed. N = 38 (WT) and 79 (Stx5ΔL) cells from 2 independent experiments. Mean ±95%CI. Not significant; unpaired two-sided Student’s t-test.

(g) Trafficking of VSVG-ts045-EGFP in parental HeLa (WT, green) or HeLa Stx5ΔL (Stx5ΔL, blue) over time after shift to 32°C (n ~40 cells/condition). When cells expressing VSVG-ts045-EGFP are cultured at 40°C, the tagged protein misfolds and is retained at the ER. Shifting the cells to 32°C at t = 0 refolds the protein and enables it to exit the ER normally and traffic to the Golgi^4^. VSVG-ts045-EGFP intensity at the Golgi was measured, corrected to the total cellular intensity of VSVG-ts045-EGFP per frame, and normalized to t = 0. See also supplementary movies 3 and 4. Shown is the mean ± error band (95%CI calculated for each timepoint and interpolated inbetween timepoints).

(h) Quantification of slope coefficients from panel (g) of the linear section (3-15 min). No differences between WT and Stx5ΔL were observed. N = 51 (WT) and 103 (Stx5ΔL) cells from 2 independent experiments. Mean ±95%CI. Not significant; unpaired two-sided Student’s t-test.

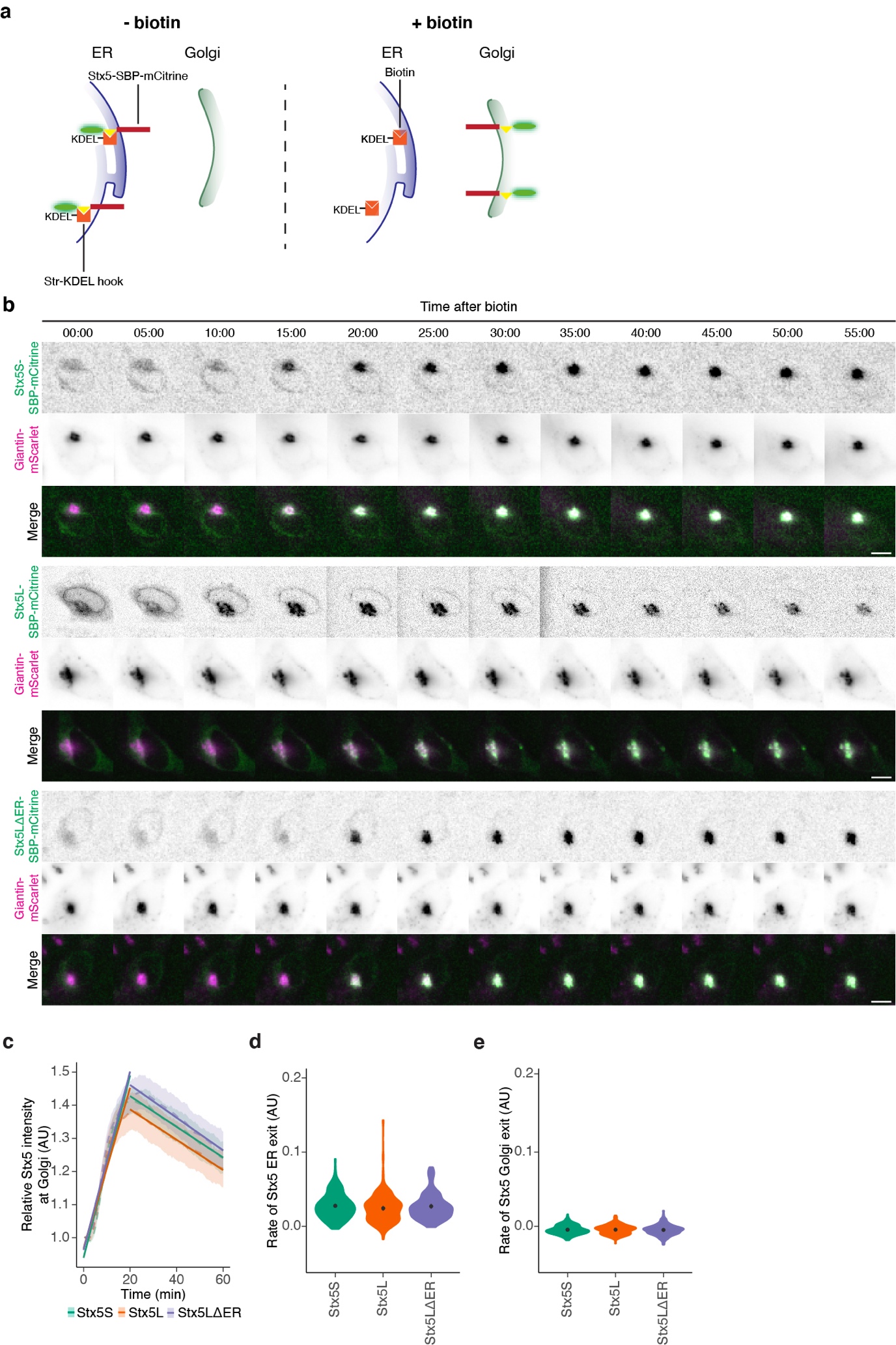

**Supplementary Figure 16. No difference in ER-Golgi trafficking of Stx5S and Stx5L isoforms.**

(a) Schematic overview of the design of Stx5 trafficking experiment, based on the RUSH system. In absence of biotin (left panel), the reporter cargo (Stx5-SBP-mCitrine) is trapped at the ER by the lumenal Str-KDEL hook. When biotin is added (right panel), biotin outcompetes the interaction with streptavidin, allowing Stx5-SBP-mCitrine to traffic freely to its destination compartment. SBP, streptavidin binding protein; Str, streptavidin.

(b) Snapshots of live-cell imaging of Stx5-SBP-mCitrine (green in merge). Magenta: Golgi marker Giantin-mScarlet. Scale bars, 10 µm. See also supplementary movies 5 – 7.

(c) Quantification of mCitrine fluorescence at the Golgi of Stx5S-SBP-mCitrine (green), Stx5L-SBP-mCitrine (orange) and Stx5LΔER-SBP-mCitrine (blue) over time from panel (b). N = 172 (Stx5S), 143 (Stx5L) and 118 (Stx5ΔER) cells from 4 independent experiments. Dashed lines: average ± error bands (95%CI for each frame interpolated between frames). Solid lines: Linear regression analysis of intensity decays of the post-ER section (0 – 20 mins) and post-Golgi section (20 – 60 mins).

(d) Quantification of the slopes from the linear regression analyses for the post-ER section of panel (c) (0 – 20 mins). Mean ± 95% CI. One-way ANOVA followed by post-hoc Tukey’s. No significant comparisons.

(e) Quantification of the slopes from the linear regression analyses of panel (c) for the Golgi exit section (20 – 60 mins). Mean ± 95% CI. One-way ANOVA followed by post-hoc Tukey’s. No significant comparisons.

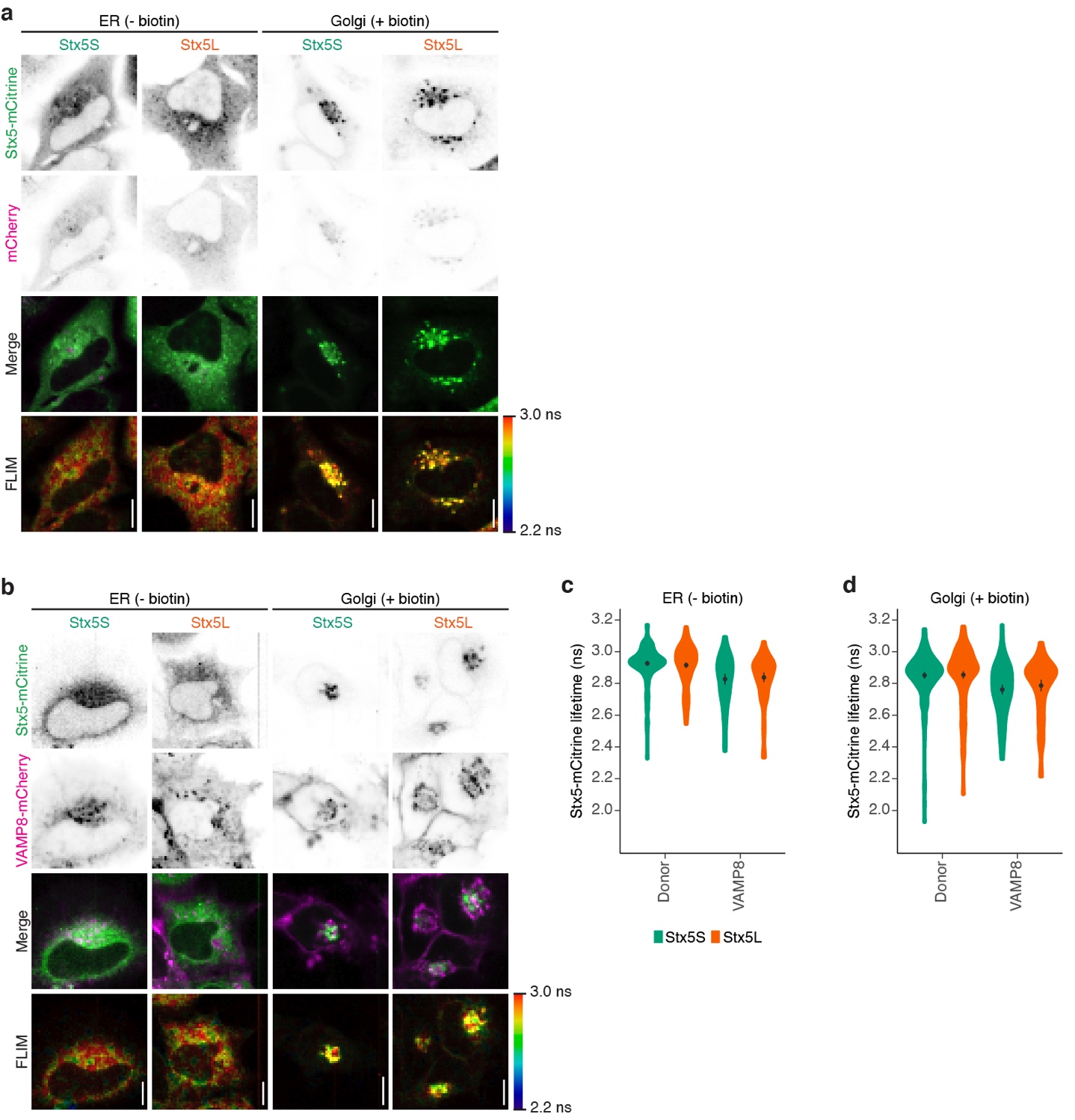
**Supplementary Figure 17. Fluorescence lifetime imaging microscopy (FLIM) of Stx5-mCitrine or with co-expression of VAMP8-mCherry.**

(a) Representative confocal micrographs and FLIM images of HeLa cells expressing Stx5S-mCitrine or Stx5L-mCitrine (green in merge) without (ER) or with (Golgi) biotin. Scalebars, 10 µm.

(b) Same as panel (a), but now for HeLa cells co-expressing Stx5-mCitrine (green in merge) and VAMP8-mCherry (magenta). Scalebars, 10 µm.

(c-d) Stx5-mCitrine lifetimes at the ER (c) and Golgi (d) from panels (a-b). Donor: donor only control with cells expressing Stx5S-SBP-mCitrine or Stx5L-SBP-mCitrine without VAMP8-mCherry. N = 52 (Stx5S Donor ER), 74 (Stx5L Donor ER), 51 (Stx5S VAMP8 ER), 61 (Stx5L VAMP8 ER), 50 (Stx5S Donor Golgi), 71 (Stx5L Donor Golgi), 39 (Stx5S VAMP8 Golgi) and 53 (Stx5L VAMP8 Golgi) cells from four independent experiments. Mean ±95%CI. Paired two-sided Student’s t-test after taking the averages for each experiment. No comparisons were significant.

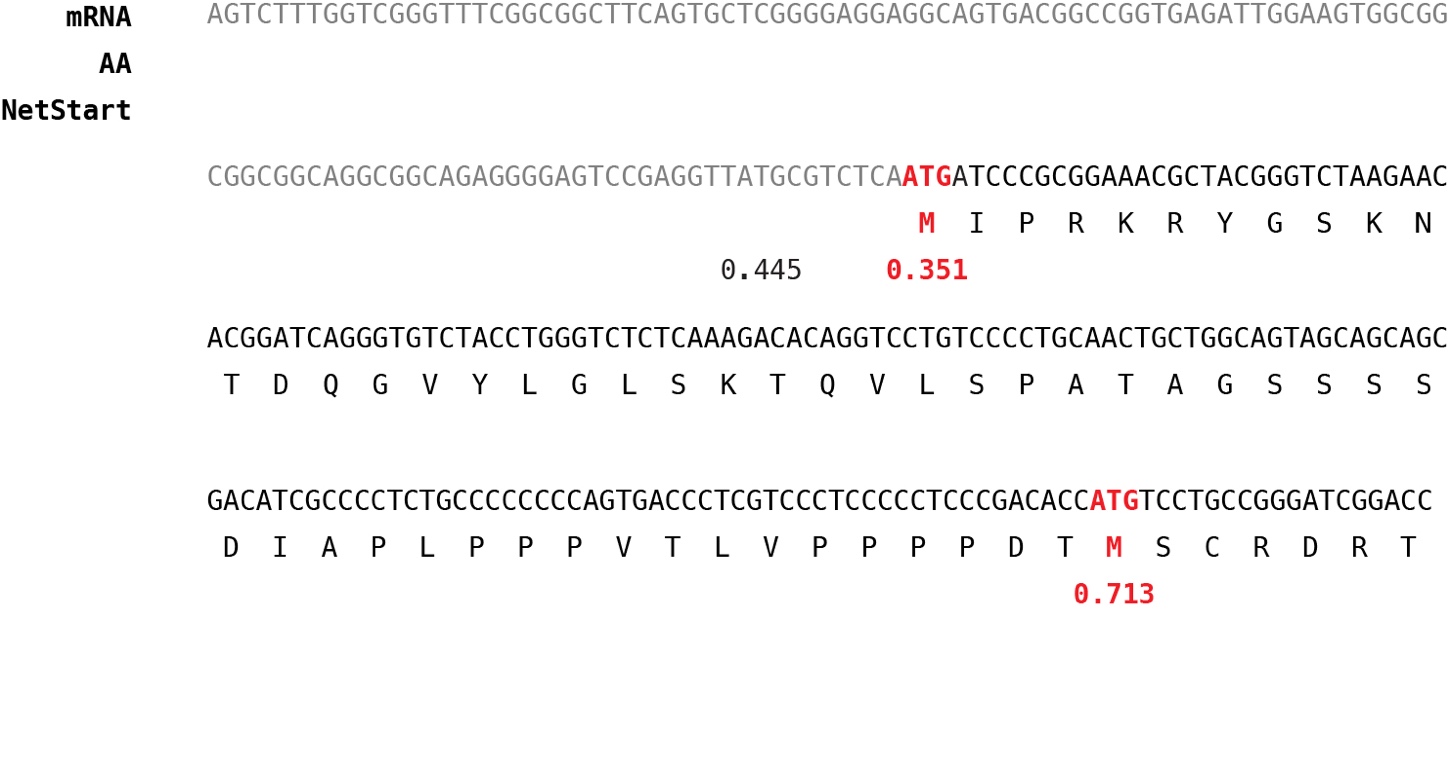

**Supplementary Figure 18. Translation initiation predictions of *STX5.***

NetStart 1.0^5^ (<https://services.healthtech.dtu.dk/service.php?NetStart-1.0>) was used to predict translation start of the *STX5* mRNA transcript (accession code: NM_003164.5 [https://www.ncbi.nlm.nih.gov/nuccore/NM_003164]). The starting codons of Stx5L and Stx5S respectively are shown in bold red. In gray is the 5’ UTR of STX5. AA, amino acid.

**Supplementary Table 1. Phenotypes of patients affected by Stx5M55V genetic variant.** Informed consent to carry out the research and publish the results was obtained from the family of investigated individuals.

|  | **Patient IV:9** | **Patient IV:10** |
| --- | --- | --- |
| Gestational age (weeks) | 29 | 37 |
| Birth weight/length/head circumference | 1380 gr/36.2 cm/29.5 cm | 2656 gr/41.5 cm/34 cm |
| Cardiovascular involvement | Ventricular septal defect | Delayed closure of ductus arteriosus, patent ductus venosus |
| Renal involvement | Agenesis of left kidney | Unilateral hydronephrosis |
| Liver involvement | Liver failure, hepatomegaly, stage 3 to 4 liver fibrosis on autopsy | Liver failure, hepatomegaly, biliary cirrhosis and nodular regenerative hyperplasia on autopsy |
| Skeletal involvement | Narrow thorax, shortening of long bones, bilateral clubfeet and flection contracture of both knees | Narrow thorax, shortening of long bones |
| Nervous system | Hypotonia | Severe hypotonia, delayed motor development, brain MRI - slight enlargement of cerebrospinal fluid spaces |
| Laboratory abnormalities | Cholestasis with increased aspartate aminotransferase, high alkaline phosphatase (1611 U/L), hyperammonaemia, coagulation abnormalities (antithrombin III 6%, and protein C 20%), hypercholesterolemia (8.5 mmol/L), hyperinsulinemic (33.5 mU/L) hypoglycemia, lower TSH (1.36 mU/L) and fT4 (8,1 pmol/L), very low IGF-I (<25 μg/L) | Cholestasis with increased transaminases, high alkaline phosphatase (2732 U/L), hyperammonaemia, coagulation abnormalities (antithrombin III 17%, protein C 25%, factor VII 46%, factor XI 21%), hypercholesterolemia (9.2 mmol/L), low ceruloplasmin (0.083 g/L), hyperinsulinemic (28 mU/L) hypoglycemia, hypokalemia, -natremia, and -phosphatemia |

**Supplementary Table 5. Chromosomal microarray analysis.**

Common LCSH areas in two patients (IV:8 and IV:9).

| **Chromosome** | **Starting position (bp) hg19** | **Ending position (bp) hg19** |
| --- | --- | --- |
| 2 | 41 051 123 | 47 273 511 |
| 5 | 113 980 360 | 123 909 500 |
| 11 | 23 410 903 | 32 583 695 |
| 11 | 33 423 590 | 48 994 066 |
| 11* | 55 091 268 | 76 733 313 |

Common LCSH areas in three patients (IV:8, IV:9 and IV:10).

| **Chromosome** | **Starting position (bp) hg19** | **Ending position (bp) hg19** |
| --- | --- | --- |
| 11 | 23 410 903 | 32 583 695 |
| 11 | 33 423 590 | 48 994 066 |
| 11* | 55 091 268 | 70 906 073 |

**STX5* gene is located on the chromosome 11 (62 806 897-62 832 088). The variant was confirmed by Sanger sequencing as homozygous in all affected individuals, and heterozygous in the mother.

**Supplementary Table 6. Antibodies used in this study.**

| **Target** | **Catalog number** | **Application** |
| --- | --- | --- |
| Stx5 (polyclonal) | Synaptic Systems, 110 053 | IF (5 µg/mL) |
| Stx5 (B-8) | Santa Cruz Biotechnology, sc-365124 | WB (0.4 µg/mL), IF (4 µg/mL) |
| Stx5L | Abcam, ab217130 | WB |
| GAPDH (14C10) | Cell Signaling Technology, 2118 | WB (0.5 µg/mL) |
| MGAT1 | Abcam, ab180578 | IF (10 µg/mL), WB (2 µg/mL) |
| MAN2A1 | Abcam, ab12277 | IF (5 µg/mL) |
| ST6GAL1 | Abcam, ab225793 | IF (20 µg/mL) |
| GALNT2 (1501421) | Biolegend, 682302 | IF (5 µg/mL) |
| B4GALT1 (GT2/36/118) | Enzo, ALX-803-339-c050 | WB (2 µg/mL) |
| GALNT3 | R&D Systems AF7174 | WB (2.9 µg/mL) |
| ST6GalNacI | Abcam ab69066 | WB (4 µg/mL) |
| GM130 (35/GM130) | BD Biosciences, 610822 | WB (0.5 µg/mL), IF (2.5 µg/mL) |
| TGN46 | BioRad, AHP500GT | WB (0.25 µg/mL), IF (0.25 µg/mL) |
| βCOP | Abcam, ab2899 | WB (4.4 µg/mL), IF (2.2 µg/mL) |
| Alpha-Tubulin (YOL1/34) | Novus Biologicals, NB100-1639 | WB (0.2 µg/mL) |
| GosR1 (1/GS28) | BD Biosciences, 611185 | WB (0.25 µg/mL) |
| GosR2 (25/GS27) | BD Biosciences, 611034 | WB (0.25 µg/mL) |
| Stx16 | Synaptic Systems, 110 162 | WB (1 µg/mL) |
| Bet1 (17) | Santa Cruz Biotechnology, sc-136390 | WB (0.2 µg/mL) |
| Bet1L (19/GS15) | BD Biosciences, 610960 | WB (1 µg/mL) |
| ERGIC53 (G1/93) | Alexis Biochemicals, 802-602-C100 | IF (10 µg/mL) |
| PDI (RL90) | Novus Biologicals, NB300-517 | IF (10 µg/mL) |
| ERGIC53 (OTI1A8) | Enzo, enz-ABS300-0100 | IF (10 µg/mL) |
| VSV-G (8G5F11) | Kerafast EB0010 | IF (20 µg/mL) |
